# Continuous Circulation of DENV-3 Genotype 3 in Delhi: A Single Hospital Based Surveillance Study

**DOI:** 10.1101/2020.04.25.20078899

**Authors:** Abhishek Padhi, Ekta Gupta, Gaurav Singh, Shama Parveen, Arshi Islam, Bansidhar Tarai

**Affiliations:** Department Of Clinical Virology, Institute Of Liver And Biliary Sciences, New Delhi; Centre For Interdisciplinary Research In Basic Sciences, Jamia Millia Islamia, New Delhi; Department Of Microbiology And Infection Control, Max Superspeciality Hospital, Saket, New Delhi

**Keywords:** Dengue, DENV 3, Genotype 3, Surveillance

## Abstract

**BACKGROUND:** Delhi is hyperendemic for dengue virus (DENV) where all the four DENV have been previously reported. A constant vigilance of circulating dengue virus serotypes is important in surveillance, since the introduction of a new variant to areas affected by pre-existing serotypes constitutes a risk factor for DHF and Dengue Shock Syndrome.

**OBJECTIVES:** This retrospective study was carried out with an objective to determine the circulating serotype and genotype of Dengue virus in acute phase blood samples of patients who reported to a tertiary liver care hospital in New Delhi during the last two years (2017 – 2018).

**METHODS:** The data of clinician-initiated testing for dengue NS1 antigen was searched in the institutional hospital information system. The serum sample of dengue NS1 antigen positive cases confirmed by ELISA (PANBIO, Gyeonggi-do, ROK) and a fever duration of less than 5 days were retrieved from the laboratory archive. The dengue virus serotyping on these sample were carried out by reverse transcriptase PCR. Sequencing and phylogenetic analysis was done for the capsid-pre membrane (CPrM) region to determine the genotype.

**RESULTS:** A total of 440 acute-phase samples were received. Twenty one(4.77%) were positive for dengue NS1 antigen with a mean age of 35.1 years and male to female ratio of 1.1:1. Eight cases (38.09%) were positive by Dengue RT-PCR and all belonged to DENV-3 serotypes. Phylogenetic tree analysis revealed DENV-3 clustered to genotype III with 100% homology with 2008 Indian subcontinent strain.

**CONCLUSION:** This study revealed that the present circulating dengue virus serotype in Delhi is DENV–3 genotype III. It is similar to previously isolated 2008 Indian subcontinent strain suggesting neither any change in serotype nor any further evolution of DENV-3. That explains the present relatively stable dengue endemicity in Delhi NCR.

## INTRODUCTION

Dengue is a vector borne acute arboviral infection widely prevalent in the tropics and sub tropics. According to the World Health Organization (WHO) estimates, 3.6 billion people reside in dengue endemic areas and 70 % of the actual burden is in Asia (1)(2).

Symptoms of dengue infection is wide ranging from subclinical disease to severe flu like symptoms, and some people develop severe dengue associated with number of complications such as severe bleeding, organ impairment and/or plasma leakage. Over the past two decades the burden of dengue cases has increased almost 15 folds (1).

Dengue infection is caused by a virus belonging to the family *Flaviviridae*, genus *Flavivirus* and consists of a single stranded positive sense RNA about 11 kb as its genome and encodes for three structural {capsid (C), pre-membrane (prM) and envelope (E)} and 7 non-structural proteins (3). Dengue virus (DENV) can be classified into four antigenically distinct serotypes (DENV-1, DENV-2, DENV-3, DENV-4) based on sequence diversity and each serotype is further grouped into various genotypes (4).

A lifelong immunity is attained after infection with a particular serotype and a cross protective immunity against other serotypes for a short period of time (5). Secondary infection by a different serotype is associated with severe dengue probably due to antibody-dependent enhancement (ADE) resulting in high viremia (6) (7).

Delhi is hyperendemic for DENV with co-circulation of all the four dengue serotypes (8). Delhi has witnessed a number of dengue epidemics in the past two decades due to rapid boom in urbanization, monsoon influenced subtropical climate, water storage practices and use of water coolers. These factors provide the mosquito vector *Aedes aegypti* a favourable environment to thrive and transmit DENV amongst the human population which at times gives rise to major outbreaks depending upon the circulating serotype and genotype of DENV (9) (10).

In the absence of an effective vaccine, virological surveillance and development of effective, locally adapted control programmes are the two most important arms in the prevention of dengue infection. Hence, there is a need of continuous monitoring of the circulating DENV type in a given population to develop such locally adapted control programmes. Furthermore most studies focus on the circulating serotypes during epidemics and the studies on circulating strains during the interepidemic periods are neglected and there can be significant variations for which public health systems are unprepared (11).

We carried out the present study targeting the *CprM* gene junction to elucidate the molecular epidemiology of the circulating strain of DENV in the population of Delhi post 2015 dengue epidemic.

## MATERIAL AND METHODS

### Patient selection and specimen collection

This was a retrospective study carried out from January 2017 to December 2018 in a tertiary care liver hospital in New Delhi, India. All consecutive patients with signs and symptoms (according to WHO guidelines) suggestive of DENV infections visiting the OPD or admitted in IPD were tested for DENV infection (12). Plasma was separated from blood samples and stored in the virology repository at -80 °C till further testing.

### DENV diagnosis

Diagnosis of DENV was done with Dengue NS1 Ag (PANBIO, Gyeonggi-do, ROK) if the sample was collected within the first 5 days of onset of clinical symptoms, and with Dengue IgM capture ELISA (PANBIO, Gyeonggi-do, ROK) if the sample was collected after 5 days of onset of clinical symptoms.

### RNA extraction and cDNA synthesis

All the DENV NS1 Ag-positive cases were further processed for RNA extraction using QIAamp Viral Mini kit (Qiagen,GmBH, Germany) according to the manufactures’ instructions. The extracted viral RNA was reverse transcribed to cDNA using Quantitect reverse transcriptase kit (QIAGEN, GmBH Germany) according to manufacturer’s instructions.

### Detection of DENV by RT-PCR

The cDNA was then amplified in the external round of RT-PCR using dengue virus consensus primers (D1: 5’ TCAATATGCTGAAACGCGCGAGAAACC G 3’; D2: 5’ TTGCACCAACAGTCAATGTCT TCA GGT TC 3’) in 25 μl reaction volume. The DNA product obtained was of 511 bp.

Subsequent amplification of cDNA was carried out with the dengue virus consensus forward primer (D1) and four dengue serotype-specific reverse primers (TS 1: 5’ CGTCTCAGTGATCCGGGGA 3’; TS 2: 5’ CGCCACAAGGGCCATGAACAG 3’; TS 3: 5’ TAACATCAT-CATGAGACAGAGC 3’; TS 4: 5’CTCTGTTGTCTTAAACAAGAGA 3’) as described by Lanciotti *et al* (13), but all the 4 serotype-specific primers were added in a single reaction mixture. A 1 : 20 dilution of the external PCR product was used in the nested PCR reaction. Dengue virus serotypes were identified by the size of the resulting DNA bands (DENV 1–482 bp, DENV 2–119 bp, DENV 3–290 bp, DENV 4–392 bp). Amplicons were resolved on 2% agarose gel and visualized with ethidium bromide in UV light by using a gel documentation system (Wealtec, USA).

### DNA Sequencing

PCR amplicons were gel-purified using QIAquick gel extraction kit (QIAGEN, GmBH Germany) as per manufacturer’s instructions. Purified amplicons were subjected to Sanger sequencing using ABI 3730 DNA analyzer (Thermofisher Scientific, MA, Waltham).

### Phylogenetic analysis

Identity of the obtained sequences were confirmed by BLAST tool available at NCBI (http://blast.ncbi.nlm.nih.gov/Blast.cgi). Both forward and reverse sequences were manually aligned and edited to resolve the nucleotide ambiguities and to obtain consensus sequence using GeneDoc v.2.7 (http://genedoc.software.informer.com/2.7/) and BioEdit v.7.2 (http://bioedit.software.informer.com/7.2/). The study sequences were aligned with other available sequences retrieved from Genbank using CLUSTAL X2 (http://www.clustal.org/clustal2/) achieved in BioEdit v.7.2 (http://bioedit.software.informer.com/7.2/). All the new sequences of DENV-3 genome were submitted to Genbank.

The phylogenetic tree was constructed for the DENV-3 sequences by Maximum Likelihood method using algorithms implemented in MEGA6 v.6.06 software. Genetic distances were calculated using Tamura-Nei model of nucleotide substitution. The robustness of the tree was assessed with 1000 bootstrap replicates.

### ETHICS

The study was approved by the Institutional ethics committee (IEC) of Institute of Liver and Biliary Sciences (ILBS) and patient consent form was waived off because of the use of leftover clinical samples sent to virology laboratory for routine diagnosis for DENV.

## RESULTS

A total of 440 acute-phase samples were received. Twenty one(4.77%) were positive for dengue NS1 antigen with a mean age of 35.1 years and male to female ratio of 1.1:1. Eight cases (38.09%) were positive by Dengue RT-PCR and all belonged to DENV-3 serotypes. No case of concomitant infection with more than one serotype was observed. In cases where NS1 antigen was positive but RT-PCR was negative, duration of fever was 5 days, as by this time viremia declines and only NS1 antigen persists till IgM antibodies are formed.

### DNA sequence analysis

The CprM region of the DENV-3 genome was sequenced for 8 positive samples. The samples were sequenced in both forward and reverse direction. All the obtained DENV-3 sequences were confirmed by BLAST. These sequences were submitted in Genbank database and accession number of the same are awaited.

### Phylogenetic analysis

Phylogenetic analysis of DENV-3 serotype was carried out for the present investigation. The H87 strain of genotype V of DENV-3 (GenBank Accession number M93130) was used as the proto-type strain. The aligned region was 240bp that corresponds to 131 to 370bp of the CprM region of full genome the prototype strain. Eighty sequences (8 studied and 72 other sequences of different genotypes of DENV-3 (GenBank) were aligned to construct Maximum Likelihood tree. Phylogenetic analysis clustered all the studied sequences with the genotype III (Indian subcontinent).

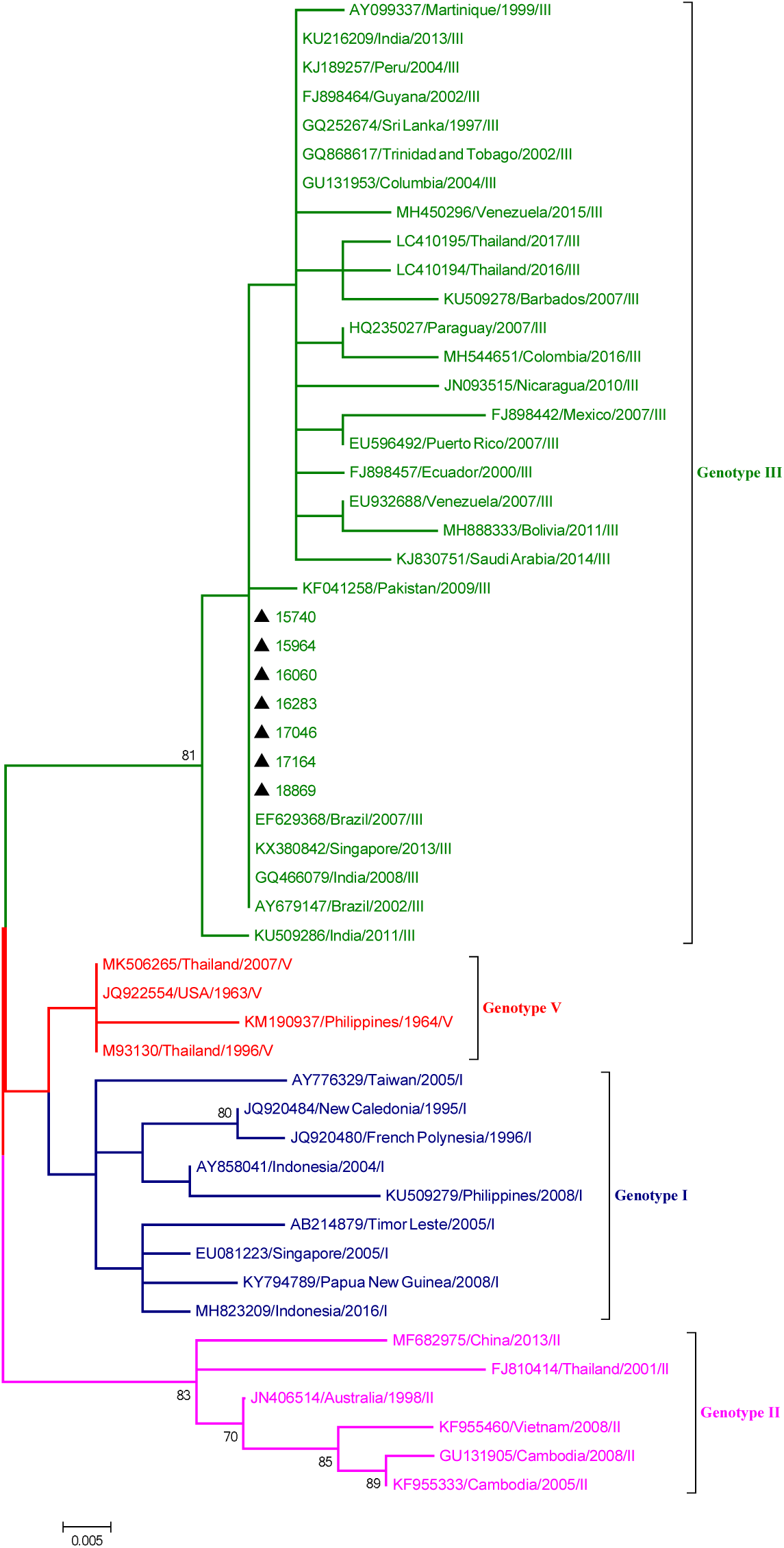
Maximum likelihood phylogenetic tree of DENV-3 strains. The study sequences are marked by the symbolΔ. Bootstrap values are represented by the number on nodes generated by 1000 replications. The strains used to construct the tree are represented by the strain name, followed by country/region and year of isolation/submission of strain. The study sequences clustered with Genotype III (Indian subcontinent).

## DISCUSSION

Delhi, the capital of India is known to be hyperendemic for DENV infection due to the circulation of all the four serotypes in the population (10). In the past two decades Delhi has witnessed several outbreaks and epidemics of DENV infection. Dense human population, rapid urbanization, increasing globalization and the tropical climatic conditions have contributed towards the frequent outbreaks of DENV infection in Delhi. Hence, a continuous sero-surveillance of the circulating DENV strain is a necessity to prevent further epidemics and help in the development of local control program (14).

In the present study, molecular characterization of circulating DENV strains was carried out targeting the *CprM* junction. Genotyping based on *CprM* junction is easier and economical because of the utilisation of a single set of primer for both amplification and sequencing of any of the four DENV serotypes (9).

A change in the circulation pattern of Dengue serotypes has been reported from Delhi in terms of prevalent serotype. The DENV-2 serotype dominated the Dengue fever during 2003 to 2006 (15)(16) (17)(18)(19). DENV-3 emerged as a dominant serotype in the year 2003 which continued in circulation till 2006 (15). In addition, 2006 also reported the emergence of DENV-1 in 30% of the cases (15). Subsequently, the predominance of DENV-1 was reported till 2010. DENV-1 and DENV-2 serotypes co-circulated in almost equal proportions in 2011 [8]. A switch in the prevailing serotype occurred again in 2012 due to co-dominant circulation of DENV-2 and DENV-3. DENV-2 dominated during 2012, 2013 and 2015 [8,15]. Subsequently, DENV-1 dominated in 2014 [16]. Further, during 2016 a change in the circulating serotype occurred leading to dominance of DENV-3 in this region.

**Table 1:** Circulation of various dengue serotypes in Delhi

| YEAR | DENV SEROTYPE | EPIDEMIC/ NON | REFERENCES |
| --- | --- | --- | --- |

Table 1 : Circulation of various dengue serotypes in Delhi
|  |  | **EPIDEMIC** |  |
| --- | --- | --- | --- |
| 1996 | 2 | Epidemic | Dar et al (20) |
| 1997 | 1 | Non epidemic | Vajpayee et al(21) |
| 2003 | 1,2,3(Genotype III),4 | Epidemic | Gupta et al (8) |
| 2004 | 1 | Non epidemic | Gupta et al(22) |
| 2005 | 3(Genotype III) | Epidemic | Gupta et al(22) |
| 2006 | 3(Genotype III) | Epidemic | Bharaj et al (23) |
| 2007 | 3(Genotype III) | Non epidemic | Kukreti et al (15) |
| 2008 | 1 | Non epidemic | Chakravarti et al (24) |
| 2010 | 1 | Non epidemic | Gupta et al (25) |
| 2011-2014 | 1,2,3(Genotype III) | Non epidemic | Afreen et al (26) |
| 2015 | 2 | Epidemic | Choudhary et al(27) |
| 2016 | 3(Genotype III) | Non epidemic | Parveen et al(28) |
| 2017-2018 | 3(Genotype III) | Non epidemic | Present study |

The most relevant finding of our study was the circulation of a single serotype, DENV-3 genotype III during 2017-2018 which was similar to the DENV-3 strain circulating in the year 2016 (28). Similar studies from Pakistan also reported the circulation of DENV-3 during 2015 (29). Studied strains of DENV-3 were clustered in Genotype III (Indian subcontinent) by phylogenetic analysis. The analysis also concluded that the DENV-3 study sequences grouped with the strains from China (unpublished), Pakistan (30) and India (30) (31) (32). Furthermore DENV-3 serotype has been reported from different geographical regions including India.

The oldest DENV-3 strain, Philippines 56, was classified as genotype I along with other strains from the Philippines and Indonesia isolated over two decades, suggesting the persistence of this genotype in the area for a long time.

It is evident that genotype III of DENV-3 circulates throughout the world, whereas other genotypes are localized in particular geographic regions. This indicates a higher potential of genotype III to spread and dominate in geographically diverse regions of the world. This genotype has also been implicated in major dengue epidemics in several parts of Asia, Africa and the Americas and has the potential to cause an international dengue pandemic (33).

Furthermore our study also shows that in the non-epidemic period usually one strain circulates and co-circulation of multiple types with concurrent infection with more than one serotype is not seen.

## CONCLUSION

The findings of this study indicate continuous circulation of DENV-3, genotype III, since 2003 in Delhi. DENV-3, Genotype III has a potential to cause global pandemic which warrants a continued surveillance to monitor its incursion and spread for effective control measures before the onset of the next outbreak. Further studies to understand the host adaptation mechanisms for vaccine implementation in our population are warranted.

## Data Availability

Data has been taken from our hospital information system

## ACKNOWLEDGEMENT

The authors would like to thank Mr. Piyush Bahuguna for his contribution in providing the data for dengue virus infection for the year 2017-2018.

